# Systematic Review and Meta-Analysis Protocol of the Efficacy and Safety of COVID-19 Drug Candidates Targeting Host Enzymes Involved in Immune Response

**DOI:** 10.1101/2023.07.28.23293338

**Authors:** Aganze Gloire-Aimé Mushebenge, Samuel Chima Ugbaja, Nonkululeko Avril Mbatha, Manimani Ghislain Riziki, Tambwe Willy Muzumbukilwa, Mukanda Gedeon kadima, Hezekiel M. Kumalo

## Abstract

**Introduction:** COVID-19 is a rapidly spreading infectious disease caused by the SARS-CoV-2 virus. Although several therapeutic interventions have been developed, the mortality rate of the disease remains high, and effective treatment options are urgently needed. Host-directed therapies targeting enzymes involved in the immune response represent a promising strategy for the development of novel therapeutics against COVID-19. This study aims to conduct a systematic review and meta-analysis of the literature to evaluate the potential of drug candidates targeting host enzymes involved in the immune response for the treatment of COVID-19.

**Methods and analysis:** We will conduct a systematic search of electronic databases including PubMed, Embase, and Cochrane Library, as well as preprint servers and clinical trial registries for relevant studies. We will include randomized controlled trials, observational studies, and preclinical studies evaluating the efficacy of drug candidates targeting host enzymes involved in the immune response in COVID-19. Two reviewers will independently screen articles, extract data, and assess study quality. The primary outcome will be the effect of drug candidates on mortality, while secondary outcomes will include time to recovery, adverse events, and changes in immune markers. A meta-analysis will be performed to estimate pooled effect sizes of the interventions, and a narrative synthesis will be conducted for studies that are not amenable to quantitative analysis. This study will provide a comprehensive evaluation of the potential of host-directed therapies targeting enzymes involved in the immune response for the treatment of COVID-19. The results of this study may guide the development of novel therapeutics against COVID-19 and inform clinical practice.

**Ethics and dissemination:** This study will review published data, and thus it is unnecessary to obtain ethical approval. The findings of this systematic review will be published in a peer-reviewed journal.

**PROSPERO registration number:** **CRD42023415110**.

## INTRODUCTION

The emergence of COVID-19 caused by the SARS-CoV-2 virus has become a global public health emergency (1). Despite the deployment of numerous interventions and the rapid development of vaccines, the pandemic is still ongoing, with new variants emerging and threatening global efforts to control the spread of the virus (2,3). While vaccination has been shown to be effective in reducing severe disease and hospitalization, there is a need for effective treatments for those who become infected, especially in areas with limited access to vaccines (4,5). Despite significant efforts to develop effective treatments, there is still a pressing need for more effective therapies to reduce the morbidity and mortality associated with this disease (6,7). The immune response to SARS-CoV-2 infection plays a critical role in determining the severity of the disease, and targeting host enzymes involved in the immune response is a promising strategy for developing novel therapeutics against COVID-19 (8,9).

The immune response to viral infection is a complex process involving multiple enzymes and pathways (10). Host-directed therapies that target enzymes involved in the immune response may provide several advantages over direct antiviral agents, including a broader spectrum of activity against different strains of viruses, reduced potential for viral resistance, and potentially fewer adverse effects (11,12). The identification of host enzymes involved in the immune response to SARS-CoV-2 infection has been the subject of intense research efforts, with several potential targets having been identified (13,14).

The pathogenesis of COVID-19 involves a complex interplay between the host immune system and the virus (15,16). During the early phase of infection, the innate immune system plays a crucial role in controlling the virus (17). However, in severe cases, a dysregulated immune response leads to hyperinflammation and cytokine storm, which can cause damage to multiple organs and ultimately result in death (18,19). Host-directed therapies targeting enzymes involved in the immune response may represent a promising strategy for the development of novel therapeutics against COVID-19.

Several host enzymes have been identified as potential targets for drug development, including cytokines, proteases, and kinases, which play crucial roles in regulating the inflammatory response and immune cell activation (15,20). For instance, cytokines such as interleukin-6 (IL-6) and tumor necrosis factor-alpha (TNF-α) are implicated in the cytokine storm associated with severe COVID-19, while proteases such as angiotensin-converting enzyme 2 (ACE2) and furin are essential for viral entry and replication (21,22).

Although several drugs targeting these enzymes are currently in clinical trials or have been approved for other diseases, there is a need for a systematic evaluation of their potential for repurposing as COVID-19 therapeutics (23,24). A systematic review and meta-analysis can provide a comprehensive evaluation of the available evidence for the efficacy and safety of these therapies. The identification of effective host-directed therapies for COVID-19 could have a significant impact on the management of this disease and reduce the burden on healthcare systems worldwide.

In this study, we aim to conduct a systematic review and meta-analysis of the literature to evaluate the potential of drug candidates targeting host enzymes involved in the immune response for the treatment of COVID-19. The findings of this study may guide the development of new therapies for COVID-19 and inform clinical practice.

## OBJECTIVES

This systematic review and meta-analysis protocol aims to explore the potential of host enzyme-targeted drugs as a novel therapeutic approach for COVID-19 treatment, by assessing their efficacy and safety compared to standard of care and other experimental treatments. The study aims to provide a comprehensive overview of the available evidence and identify knowledge gaps and research priorities in this emerging field.

### Review questions

What is the efficacy and safety of COVID-19 drug candidates targeting host enzymes involved in the immune response, compared to standard of care and other experimental treatments, in reducing the severity of COVID-19 symptoms and improving clinical outcomes?

To address this primary research question, the following sub-questions will be considered:

1. Which enzymes have been targeted for drug development and what is the evidence supporting their role in COVID-19 pathogenesis?
2. What is the clinical efficacy of drugs targeting host enzymes in reducing the severity of COVID-19 symptoms and mortality?
3. What is the safety profile of drugs targeting host enzymes, including potential adverse events and drug-drug interactions?
4. What is the efficacy of host-directed therapies targeting enzymes involved in the immune response for the treatment of COVID-19?
5. What are the potential mechanisms of action of these host-directed therapies, and how do they relate to the pathogenesis of COVID-19?

## METHODS AND ANALYSIS

The meta-analysis protocol will adhere to the Preferred Reporting Items for Systematic Review and Meta-Analysis Protocols (PRISMA-P) guidelines, which are widely recognized as the standard for transparent reporting of systematic review protocols (25). The protocol has been registered with the International Prospective Register of Systematic Reviews (PROSPERO), which is a publicly accessible database of systematic review and meta-analysis protocols, to ensure transparency and accountability in the review process. The registration number will be included in the final publication of the systematic review and meta-analysis.

### Eligibility criteria

The systematic review and meta-analysis of COVID-19 drug candidates targeting host enzymes involved in the immune response will include randomized controlled trials, non-randomized controlled trials, cohort studies, case-control studies, and case series that evaluate the efficacy and safety of host enzyme-targeted drugs for COVID-19 treatment. Studies must compare host enzyme-targeted drugs to standard of care or other experimental treatments for COVID-19, report outcomes related to the severity of COVID-19 symptoms and clinical outcomes, such as mortality, hospitalization rates, and length of hospital stay. Only studies published in English or with English translations available will be considered.

### Patients, intervention, comparison, outcome strategy and types of studies

➢ Population: Patients with confirmed COVID-19 diagnosis
➢ Intervention: Host enzyme-targeted drugs for COVID-19 treatment
➢ Comparison: Standard of care or other experimental treatments for COVID-19
➢ Outcome: Severity of COVID-19 symptoms and clinical outcomes (mortality, hospitalization rates, length of hospital stay, etc.)
➢ Study design: Randomized controlled trials, non-randomized controlled trials, cohort studies, case-control studies, and case series.

### Information sources

The following databases will be searched for the systematic review and meta-analysis of COVID-19 drug candidates targeting host enzymes involved in the immune response: PubMed, Embase, Cochrane Library, WHO COVID-19 database, medRxiv (preprint server), bioRxiv (preprint server), ClinicalTrials.gov (clinical trial registry), WHO International Clinical Trials Registry Platform (ICTRP). In addition, reference lists of relevant articles, reviews, and meta-analyses will be screened for potentially eligible studies. Grey literature, such as conference abstracts, dissertations, and government reports, will also be searched to identify unpublished or ongoing studies. Expert consultation may be sought to identify any additional relevant studies.

### Patient and public involvement

The study will not include individual patient data, and a systematic literature search will be conducted using specified databases. There will be no patient involvement in the study design, implementation, or dissemination of findings.

### Search Strategy

The search strategy for the systematic review and meta-analysis of COVID-19 drug candidates targeting host enzymes involved in the immune response will include a combination of MeSH terms and keywords related to COVID-19, host enzymes, and drug treatments. The following search strategy may be used for the PubMed database: ((COVID-19[Title/Abstract] OR SARS-CoV-2[Title/Abstract]) AND (host enzymes[Title/Abstract] OR immune response[Title/Abstract] OR inflammation[Title/Abstract] OR cytokines[Title/Abstract])) AND (drug therapy[Title/Abstract] OR pharmacology[Title/Abstract] OR therapeutics[Title/Abstract]) AND (randomized controlled trial[Publication Type] OR controlled clinical trial[Publication Type] OR cohort study[Publication Type] OR case-control study[Publication Type] OR case series[Publication Type]).

Similar search strategies will be adapted for other databases. The search will be limited to English language publications from January 2020 to the present. The reference lists of relevant articles, reviews, and meta-analyses will be screened for additional eligible studies. Grey literature sources, such as conference proceedings and dissertations, will also be searched to identify unpublished or ongoing studies. The search strategy will be documented in detail and reported following the PRISMA guidelines.

### Study records

This systematic review and meta-analysis will include randomized controlled trials, non-randomized controlled trials, cohort studies, case-control studies, and case series that evaluate the efficacy and safety of host enzyme-targeted drugs for COVID-19 treatment. Studies must report outcomes related to the severity of COVID-19 symptoms and clinical outcomes, such as mortality, hospitalization rates, and length of hospital stay. Only studies published in English or with English translations available will be considered. We will exclude studies that do not meet the inclusion criteria. The selection process will be summarized using a PRISMA flow diagram. The study selection process will be conducted by a team of four researchers who will independently screened the titles and abstracts of identified studies, excluding any duplicate studies. The same authors willl then review the full texts of potentially eligible studies and assessed their compliance with the inclusion criteria. Any discrepancies will be resolved by a fifth reviewer. A visual representation of the study selection process is presented in the flow chart below.

All details of the screening and data extraction process will be reported in Supplementary Table.

**Figure 1:**
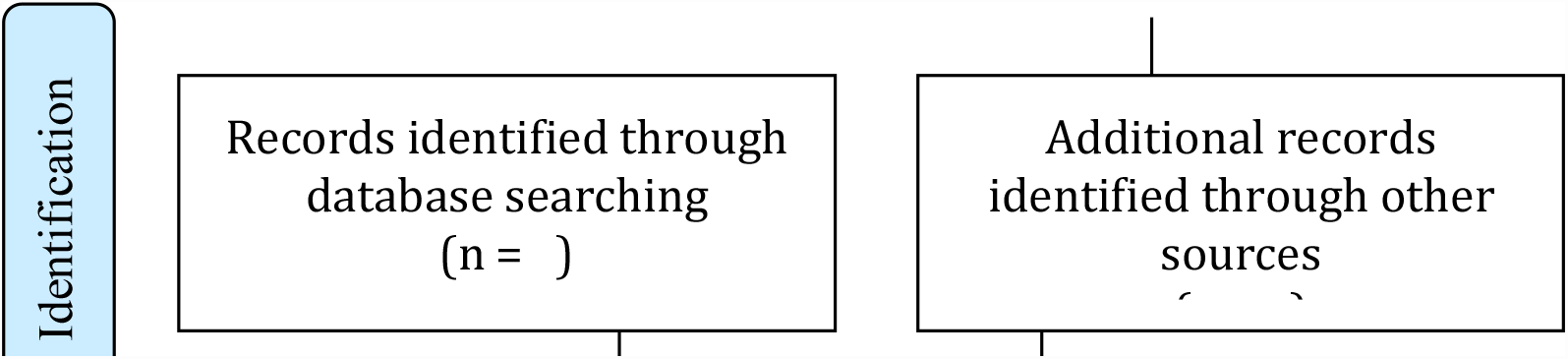
The PRISMA Flow Diagram for the systematic review screening process

### Data Collection Process and Management

A systematic search of the selected databases will be conducted, and the identified studies will be screened for eligibility by two independent reviewers. Data extraction will be performed by two reviewers using a pre-defined data extraction form. Any discrepancies will be resolved by discussion or consultation with a third reviewer. The extracted data will be entered into a Microsoft Excel spreadsheet and checked for accuracy. The data will be analysed using appropriate statistical methods and synthesized to determine the efficacy and safety of host enzyme-targeted drugs for COVID-19 treatment.

### Risk of bias in individual studies

The risk of bias in individual studies will be assessed using the Cochrane Risk of Bias tool for randomized controlled trials, the Newcastle-Ottawa Scale for non-randomized controlled trials and cohort studies, and the Joanna Briggs Institute critical appraisal checklist for case-control studies and case series (26,27). The assessment will be performed independently by two reviewers, and any discrepancies will be resolved by discussion or consultation with a third reviewer. The results of the risk of bias assessment will be reported in the systematic review and meta-analysis.

The Review Manager software (RevMan V.5.2.3) will be used to enter data, protocols, characteristics of studies, comparison tables, and perform meta-analyses. For dichotomous outcomes, the odds ratio (OR) and 95% confidence interval (CI) will be extracted or calculated for each study. In cases of high heterogeneity (I2 ≥50%), a random-effects model will be applied to combine the studies and calculate the OR and 95% CI using the DerSimonian-Laird algorithm.

### Data synthesis and analysis

#### Metabias

The Grading of Recommendations, Assessment, Development and Evaluation (GRADE) approach will be utilized to evaluate the strength of evidence derived from the included data (28). The assessment summary will be integrated with other measurements to ensure the evaluation of risk of bias, consistency, directness, and precision (28,29). The quality of evidence will be evaluated based on factors such as risk of bias, indirectness, inconsistency, imprecision, and publication bias (29).

Potential publication bias will be assessed using funnel plots and Egger’s test. In addition, we will conduct a sensitivity analysis by excluding studies with a high risk of bias to evaluate the impact of bias on the overall results (30). We will also perform subgroup analyses based on study characteristics such as study design, sample size, and risk of bias to assess the impact of these factors on the overall findings. The results of the meta bias assessment will be reported in the systematic review and meta-analysis.

#### Strengths and limitations

One of the strengths is the comprehensive search strategy that will be employed, which includes multiple databases and a thorough screening process. Additionally, the use of pre-specified inclusion and exclusion criteria will ensure that the studies selected for inclusion are relevant to the research question.

Another strength is the use of the GRADE approach to assess the quality of evidence. This will provide a transparent and rigorous assessment of the strength of the evidence and will allow for a more informed interpretation of the results.

However, there are also several limitations to consider. One potential limitation is the reliance on published studies, which may introduce publication bias and may not capture all relevant data. Additionally, the inclusion of only studies published in English may exclude relevant studies in other languages.

Another potential limitation is the heterogeneity of the included studies, which may make it difficult to draw meaningful conclusions from the meta-analysis. Finally, the quality of the studies included in the meta-analysis may vary, which could affect the overall quality of the evidence.

Despite these limitations, this systematic review and meta-analysis protocol represents an important step in synthesizing the current evidence on the efficacy and safety of COVID-19 drug candidates targeting host enzymes involved in immune response.

## DISCUSSION

The ongoing COVID-19 pandemic has posed a significant challenge to the healthcare system worldwide, and several drug candidates targeting host enzymes involved in immune response have been proposed for the treatment of this disease. This systematic review and meta-analysis protocol aims to provide a comprehensive evaluation of the efficacy and safety of these drug candidates.

This review protocol follows the Preferred Reporting Items for Systematic Review and Meta-Analysis Protocols (PRISMA-P) guidelines to ensure the highest standards of reporting. The eligibility criteria have been carefully designed to select studies that meet the objectives of this review, and a comprehensive search strategy will be used to identify relevant studies.

The data collection process and management plan will ensure the accuracy and reliability of the results. The risk of bias in individual studies will be assessed using the Cochrane Risk of Bias tool, and the strength of evidence will be evaluated using the GRADE approach. In addition, we will assess potential publication bias using funnel plots and Egger’s test and perform sensitivity and subgroup analyses to assess the impact of bias and other factors on the overall findings.

The findings of this study will have significant implications for the clinical management of COVID-19 patients. The results will provide valuable insights into the safety and efficacy of the proposed drug candidates and help guide clinical decision-making. Additionally, the study will identify areas where further research is needed to improve our understanding of the treatment of COVID-19.

It is important to note that this study has some limitations. Firstly, the search will be limited to English language publications, potentially introducing language bias. Secondly, the inclusion of only published studies may introduce publication bias. Finally, the quality of the included studies may vary, potentially affecting the strength of evidence. However, these limitations will be acknowledged and addressed through sensitivity analyses and the GRADE approach.

In conclusion, this systematic review and meta-analysis protocol will provide a comprehensive evaluation of the efficacy and safety of COVID-19 drug candidates targeting host enzymes involved in immune response. The study will provide valuable insights into the clinical management of COVID-19 patients and identify areas where further research is needed.

## Data Availability

All data produced in the present work are contained in the manuscript

## Ethics and dissemination

This study will review published data, and thus it is unnecessary to obtain ethical approval. The findings of this systematic review will be published in a peer-reviewed. journal.

## Contributors

AGAM and HMK contributed to the design of this review. AGAM drafted the protocol manuscript. AGAM, SCU and NAM revised the manuscript. MGR, TWM and MGK developed the search strategies. AGAM and SCU implemented the search strategies. AGAM, SCU and NAM tracked the potential studies, extracted the data, and assessed the quality. In case of disagreement between the data extractors, MN advised on the methodology and worked as a referee. AGAM and HMK completed the data synthesis. All authors approved the final version for publication.

## Patient and public involvement

Patients and/or the public were not involved in the design, or conduct, or reporting, or dissemination plans of this research.

## Patient consent for publication

Not applicable.

